# A Three-Item History-Based Model to Estimate Fatigue Severity: A Large Cross-Sectional Community Study across Five Neurological Conditions

**DOI:** 10.64898/2025.12.19.25342360

**Authors:** Patricia Fuller, Sarah Fearn, Benjamin Barton, Sandra Bartolomeu Pires, Veena Agarwal, Christopher Kipps

## Abstract

**Background:** Fatigue is prevalent and debilitating across neurological disorders yet remains under recognised in routine care. We characterised and compared fatigue across five long term neurological conditions using a common assessment, and developed a brief, history-based model to estimate fatigue severity.

**Methods:** Adults with epilepsy, Huntington’s disease (HD), multiple sclerosis (MS), Parkinson’s disease (PD), or motor neurone disease (MND) completed an online cross-sectional survey (n=652). The outcome was self-rated fatigue severity (1-10 numerical rating scale). The development sample comprised complete cases (n=481); multiple imputation sensitivity analyses (m=20) were undertaken. Model performance was assessed with adjusted R2, root mean squared error (RMSE), mean absolute error (MAE), and calibration (slope/intercept) with bootstrap resampling and 10-fold cross-validation (CV).

**Results:** Clinically significant fatigue (>=6/10) affected 51.1% of participants. Highest rates were in MND and MS, and lowest in PD. Fatigue episodes occurred at least twice weekly in 70.4% of participants (epilepsy 60.1%, HD 58.3%, MS 79.0%, PD 69.2%, MND 82.3%). Fatigue impacted social life and concentration “often” or more frequently in 38.5% and 34.5% of participants, respectively. Overall, 69.8% had not discussed fatigue with a healthcare professional. In multivariable models adjusted for demographics and diagnosis, three patient-reported items independently predicted severity: episode frequency, frequency of impact on social life, and frequency of impact on concentration. Full model performance: CV adjusted R2=0.45; RMSE=1.59; MAE=1.27; slope=0.92 (0.84-1.01), intercept=+0.39 (−0.06 to +0.91). A three-item history-based model performed similarly (CV adjusted R2=0.47; RMSE=1.57; MAE=1.25; slope=0.96 (0.87-1.05); intercept=+0.21 (−0.30 to +0.73). Diagnosis was not independently associated with severity after adjustment; episode frequency was the strongest predictor.

**Conclusions:** Fatigue is frequent and functionally disruptive across neurological conditions, yet seldom discussed in clinic. We derived a three-item history-based model that explained nearly half of the variance in self-rated severity, showing stable internal performance with good calibration. Because episode frequency and frequency of impact are rarely captured by existing fatigue scales, eliciting them as simple history items offers a pragmatic, transdiagnostic triage approach for clinical and digital pathways. External validation across neurological and other long-term conditions is now required.

## Background

Fatigue is one of the most distressing and disabling symptoms experienced by people living with long-term neurological conditions. It is often described as an overwhelming, persistent exhaustion that is disproportionate to effort and unrelieved by rest. However, despite its impact on quality of life and recognition in national guidelines [1, 2], fatigue remains poorly understood and frequently under-recognised across neurological care [1–8].

Clinically significant fatigue, identified using validated self-report instruments, is highly prevalent. Recent systematic reviews report prevalence of 50% in Parkinson’s disease [3], 47% in epilepsy [4] and up to 78% in MS [5]. Smaller studies in Huntington’s disease and motor neurone disease report estimates ranging from 29% to 83%, with estimates varying depending on disease stage and sample characteristics [6–14].

However, much less is known about how often episodes of fatigue are experienced. Most studies in MS were conducted before contemporary diagnostic classifications. A recent survey found that 84.1% of people with MS reported fatigue ‘often’ or ‘always’ [15]. In PD, only a single study has explored episode frequency, reporting that 67% of participants experienced fatigue at least twice weekly [16]. Equivalent data are lacking for epilepsy, MND, and HD, highlighting the limited understanding of temporal fatigue patterns.

With respect to functional impact, higher fatigue severity in MS is associated with poorer social functioning [17–24], with up to 90⍰% of patients with progressive disease reporting social limitations [15, 25]. Sub-scale data in PD and epilepsy demonstrate similar associations [26–28], though data for MND and HD remain sparse. In MS [29–31], PD [32, 33], and epilepsy [34] fatigue has also been linked to slower processing speed and reduced attention, though findings have not always been consistent [35].

These gaps have clinical implications. Fatigue has been linked to relapse, crisis events, and escalating care needs [36] and emergency care use in neurological populations is nearly double that of other long-term conditions [37]. Yet, available fatigue tools are often time-intensive, condition-specific, and focus primarily on severity [38].

In response, we conducted a large, community-based survey of individuals living with five long-term neurological conditions. Epilepsy, Huntington’s disease, multiple sclerosis, Parkinson’s disease, and motor neurone disease were selected to represent the heterogeneity across neurological practice, demographics, and disease trajectory. Our aims were twofold: (1) to characterise fatigue across conditions using a common measure to allow valid comparison, and (2) to develop a brief, clinically usable model to estimate fatigue severity from patient-reported history alone.

## Methods

### Study Design

This was a cross-sectional, community-based survey of adults living in England with epilepsy, HD, MS, PD, and MND. Recruitment took place between 2018 and 2021 via National Health Service (NHS) outpatient clinics in Southern England and through national and local support groups.

Eligibility criteria were age ≥18 years, confirmed clinical diagnosis of one of the above conditions, with capacity to give informed consent.

The survey was hosted on a secure university platform. Of 767 individuals who provided informed consent, 115 returned empty or single-item responses and were excluded, yielding 652 surveys for analysis. The complete-case sample for model development comprised 481 participants.

### Measures

#### Outcome

The primary outcome was self-rated fatigue severity over the past month, assessed on a 1 to 10 numerical rating scale (NRS). The scale was chosen due to its brevity, ease of use and prior validation in neurology populations. Fatigue was defined as ‘intense or severe tiredness or exhaustion’.

#### Candidate predictors

Prespecified predictors were collected, adapted from a survey initially developed to explore fatigue in PD (see Supplementary files for full questionnaire [16]). These included:

- **Sociodemographic variables:** age group, gender, living status (alone vs not), and working status (working vs retired).
- **Clinical variables:** neurological condition, comorbidity count (0, 1, 2, ≥3), and duration since diagnosis (years).
- **Fatigue descriptors:** frequency of fatigue episodes; frequency of functional disruption to social life, movement and concentration; impact on daily activities, exercise, hobbies, falls/balance; predictability and fluctuation; perceived change since diagnosis; agreement with descriptors used for fatigue.

#### Patient and public involvement

People with lived experience of neurological conditions reviewed the survey wording, burden, and accessibility.

### Statistical analysis

Analyses used Stata 18 (StataCorp LLC, College Station, TX, USA). Two-sided p<0.05 was considered statistically significant; estimates are reported with 95% confidence intervals (CI). Reporting follows TRIPOD recommendations [39].

#### Descriptive analysis

Categorical variables were summarised as n (%); continuous variables as mean (SD) and median (IQR). Fatigue severity and descriptors were reported overall and by condition. For descriptive and illustrative purposes, fatigue severity was also grouped into three prespecified bands: low (<4.0), moderate (4.0–5.9), and high (≥6.0). These thresholds were chosen as pragmatic interpretive categories, consistent with conventions for defining clinically significant fatigue [40, 41] and symptom rating [42, 43] rather than as empirically derived cut-points.

#### Model objective and intended use

The primary objective was to derive a brief, interpretable, history-based estimator of fatigue severity. The intended use was early recognition and triage in clinical and digital pathways, with emphasis on interpretability and calibration.

#### Model specification

Multivariable linear regression models were fitted with fatigue severity (1 to 10 NRS) as the dependent variable. Linear regression was preferred over ordinal approaches for simplicity of interpretation and comparability with other symptom scales; ordinal sensitivity analyses confirmed similar graded relationships and the same three history items as dominant predictors (see Supplementary files). The following a priori covariates were forced into all models to support face validity and potential transportability across clinics and case-mixes: age group, sex, neurological diagnosis, comorbidity count, living status, working status, and duration since diagnosis. Categorical predictors were dummy coded with the following reference categories: condition (PD), age (≥65 years), gender (female), living status (not alone), working status (working), comorbidity count (0), and ‘Never’ for frequency/impact items. Model assumptions (linearity, collinearity, normality and homoscedasticity of residuals) were assessed using standard diagnostics; no important violations were identified.

#### Item reduction and parsimony

To obtain a parsimonious, clinically usable model, we applied structured item reduction. Candidate fatigue history items were first inspected in univariable models and then entered into multivariable models, retaining items that improved adjusted R^2^ and clinical interpretability. Forced covariates (age, sex, diagnosis, comorbidity, living/working status, duration since diagnosis) were retained in all full models. To address potential optimism from data-driven item reduction, we used bootstrap resampling (500 bootstrap resamples) for coefficient uncertainty and 10-fold cross-validation for performance and calibration.

#### Missing data

The primary analysis used complete cases (n=481/652). Missingness per variable was 3 to 8% and dispersed across many small patterns. As a sensitivity analysis under a Missing At Random assumption, multiple imputation by chained equations (m=20) was performed using appropriate models for each variable type, including all analysis variables and the outcome. The regression model was re-estimated in the imputed dataset with robust standard errors, and direction and magnitude of coefficients were compared with the complete-case model. Characteristics of included versus excluded participants were compared using χ^2^ tests.

#### Internal validation and calibration

Model performance was summarised using adjusted R^2^, root mean squared error (RMSE), mean absolute error (MAE), and calibration (slope and intercept). Internal validation was performed using 10 fold cross-validation to obtain out-of-fold predictions for performance metrics and calibration.

#### Sample size

The final analytic sample (n=481) yielded approximately 15.5 observations per model degree of freedom (31 df), exceeding common rules of thumb for linear prediction model development which recommend a minimum of 10 to 20 observations per predictor parameter [44].

## Results

### Sample characteristics

A total of 652 participants were included (Table 1). There were slightly more female than male participants overall, with women predominating in MS and epilepsy, and men in PD, HD and MND. The modal age group was ≥65 years, reflecting the large proportion of PD (48%) in the cohort. Most participants (75.8%) reported one or no comorbidities, with HD having the highest proportion reporting none. Median disease duration varied by condition, from 1.9 years in MND to 11.8 years in epilepsy, consistent with typical disease trajectories. Most were retired and not living alone, although higher proportions of MS and epilepsy participants remained in work, reflecting their younger age profile.

**Table 1.** Demographic characteristics for the whole sample and stratified for each neurological condition.

| Variable | Group | All | Epilepsy | Huntington's Disease | Multiple Sclerosis | Parkinson's Disease | Motor Neurone Disease |
| --- | --- | --- | --- | --- | --- | --- | --- |
| Number (%) |  | 652 | 123 (18.9) | 12 (1.8) | 166 (25.5) | 317 (48.6) | 34 (5.2) |
| Age | 18-34 | 45 (6.9) | 42 (34.1) | 0 | 3 (1.8) | 0 | 0 |
|  | 35-44 | 42 (6.4) | 12 (9.8) | 0 | 28 (16.9) | 2 (6.0) | 0 |
|  | 45-54 | 97 (14.9) | 23 (18.7) | 4 (33.3) | 53 (31.9) | 13 (4.1) | 4 (11.8) |
|  | 55-64 | 126 (19.3) | 19 (15.4) | 2 (16.7) | 46 (27.7) | 50 (15.8) | 9 (26.5) |
|  | ≥65 | 300 (46.0) | 22 (17.9) | 4 (33.3) | 29 (17.5) | 233 (73.5) | 12 (35.3) |
|  | Missing | 42 (6.4) | 5 (4.1) | 2 (16.7) | 7 (4.2) | 19 (5.7) | 9 (26.5) |
| Gender | Female | 346 (53.1) | 77 (62.6) | 4 (33.3) | 119 (71.7) | 132 (41.6) | 14 (41.2) |
|  | Male | 288 (44.2) | 41 (33.3) | 7 (58.3) | 43 (25.9) | 179 (56.5) | 18 (52.9) |
|  | Missing | 18 (2.8) | 5 (4.1) | 1 (8.3) | 4 (2.4) | 6 (1.9) | 2 (5.9) |
| Live alone | Yes | 124 (19.0) | 22 (17.9) | 4 (33.3) | 25 (15.1) | 67 (21.1) | 6 (17.6) |
|  | No | 513 (78.7) | 99 (80.5) | 7 (58.3) | 138 (84.7) | 245 (77.3) | 24 (70.6) |
|  | Missing | 15 (2.3) | 2 (1.6) | 1 (8.3) | 3 (1.8) | 5 (1.6) | 4 (11.8) |
| Working status | Retired | 408 (62.6) | 38 (30.9) | 9 (75.0) | 61 (36.7) | 272 (85.8) | 28 (82.4) |
|  | Working | 203 (31.1) | 68 (55.3) | 2 (16.7) | 90 (54.2) | 39 (12.3) | 4 (11.8) |
|  | Missing | 41 (6.3) | 17 (13.8) | 1 (8.3) | 15 (9.0) | 6 (1.9) | 2 (5.9) |
| Duration since diagnosis (Yrs) | Median | 6.5 | 11.8 | 4.3 | 11.4 | 5.1 | 1.9 |
|  | IQR | 2.6-13.0 | 3.4-31 | 3.8-13.8 | 5.6-20.5 | 2.1-8.5 | 0.8-8.4 |
|  | Missing | 37 (5.7) | 10 (8.1) | 1 (8.3) | 8 (4.8) | 15 (4.7) | 3 (8.8) |
| Comorbidities | None | 321 (49.2) | 59 (48.0) | 10 (83.3) | 97 (58.4) | 133 (42.0) | 22 (64.7) |
|  | One | 174 (26.6) | 31 (25.2) | 2 (16.7) | 31 (18.7) | 103 (32.5) | 7 (20.6) |
|  | Two | 94 (14.4) | 23 (18.7) | 0 | 21 (12.7) | 45 (14.2) | 5 (14.7) |
|  | ≥ Three | 56 (8.6) | 10 (8.1) | 0 | 14 (8.4) | 32 (10.1) | 0 |
|  | Missing | 7 (1.2) | 0 | 0 | 3 (1.8) | 4 (1.3) | 0 |

### Frequency of fatigue episodes

Episodes of fatigue were common, with 70.4% of participants reporting episodes two or more times per week (Table 2). This was highest in MND (82.3%) and MS (79.0%), followed by PD (69.2%), epilepsy (60.1%), and HD (58.3%). Conversely, epilepsy had the highest proportion reporting fatigue only once a week or less (35.8%). Sex differences were also observed: men with MND reported greater fatigue frequency than women, while women with MS reported greater fatigue frequency than men (Figure 1).

**Table 2.** Characteristics of fatigue for the whole sample and stratified by neurological condition.

| Characteristics | Subgroup | All | Epilepsy<br>N=123 | HD<br>N=12 | MS<br>N=166 | PD<br>N=317 | MND<br>N=34 |
| --- | --- | --- | --- | --- | --- | --- | --- |
| Number (%) |  |  |  |  |  |  |  |
| How often have you felt fatigued in the last month? | Every day, all the time | 66 (10.1) | 11 (8.9) | 1 (8.3) | 23 (13.9) | 23 (7.3) | 8 (23.5) |
|  | Almost every day for a few hours | 242 (37.1) | 31 (25.2) | 5 (41.7) | 65 (39.2) | 127 (40.1) | 14 (41.2) |
|  | 2-5 times per week | 151 (23.2) | 32 (26.0) | 1 (8.3) | 43 (25.9) | 69 (21.8) | 6 (17.6) |
|  | Once a week | 56 (8.6) | 8 (6.5) | 0 | 9 (5.4) | 38 (12.0) | 1 (2.9) |
|  | Once a fortnight | 36 (5.5) | 12 (9.8) | 0 | 9 (5.4) | 15 (4.7) | 0 |
|  | Once a month | 30 (4.6) | 9 (7.3) | 1 (8.3) | 5 (3.0) | 15 (4.7) | 0 |
|  | Never | 49 (7.5) | 15 (12.2) | 1 (8.3) | 10 (6.0) | 20 (6.3) | 3 (8.8) |
|  | Missing | 22 (3.4) | 5 (4.1) | 3 (25.0) | 2 (1.2) | 10 (3.2) | 2 (5.9) |
| Does fatigue affect your social life? | All the time | 60 (9.2) | 11 (8.9) | 1 (8.3) | 21 (12.7) | 21 (6.6) | 6 (17.6) |
|  | Most of the time | 84 (12.9) | 14 (11.4) | 1 (8.3) | 26 (15.7) | 37 (11.7) | 6 (17.6) |
|  | Often | 107 (16.4) | 17 (13.8) | 1 (8.3) | 34 (20.5) | 47 (14.8) | 8 (23.5) |
|  | Sometimes | 201 (30.8) | 37 (30.1) | 3 (25.0) | 50 (30.1) | 105 (33.1) | 6 (17.6) |
|  | Rarely | 90 (13.8) | 16 (13.0) | 1 (8.3) | 15 (9.0) | 54 (17.0) | 4 (11.8) |
|  | Never | 59 (9.0) | 18 (14.6) | 2 (16.7) | 12 (7.2) | 26 (8.2) | 1 (2.9) |
|  | Missing | 51 (7.8) | 10 (8.1) | 3 (25.0) | 8 (4.8) | 27 (8.5) | 3 (8.9) |
| Does fatigue affect your concentration? | All the time | 51 (7.8) | 16 (13.0) | 2 (16.7) | 13 (7.8) | 16 (5.0) | 4 (11.8) |
|  | Most of the time | 66 (10.1) | 13 (10.6) | 1 (8.3) | 24 (14.5) | 26 (8.2) | 2 (5.9) |
|  | Often | 108 (16.6) | 22 (17.9) | 1 (8.3) | 36 (21.7) | 44 (13.9) | 5 (14.7) |
|  | Sometimes | 200 (30.7) | 41 (33.3) | 1 (8.3) | 51 (30.7) | 99 (31.2) | 8 (23.5) |
|  | Rarely | 121 (18.6) | 8 (6.5) | 3 (25.0) | 25 (15.1) | 77 (24.3) | 8 (23.5) |
|  | Never | 63 (9.7) | 14 (11.4) | 2 (16.7) | 10 (6.0) | 34 (10.7) | 3 (8.8) |
|  | Missing | 43 (6.6) | 9 (7.3) | 2 (16.7) | 7 (4.2) | 21 (6.6) | 4 (11.8) |
| **Have you sought advice from a health care professional?** | Yes | 182 (27.9) | 22 (17.9) | 4 (33.3) | 82 (49.4) | 61 (19.2) | 13 (38.2) |
|  | No | 421 (64.6) | 87 (70.7) | 7 (58.3) | 76 (45.8) | 233 (73.5) | 18 (52.9) |
|  | Missing | 49 (7.5) | 14 (11.4) | 1 (8.3) | 8 (4.8) | 23 (7.3) | 3 (8.8) |
| **Rate your average levels of tiredness over the last month from 1-10** | Mean | 5.55 | 5.48 | 5.50 | 5.91 | 5.34 | 5.97 |
|  | Standard deviation | 2.17 | 2.50 | 2.15 | 2.10 | 2.06 | 2.14 |
|  | Median | 6 | 6 | 6 | 6 | 5 | 6 |
|  | IQR | 4-7 | 3-8 | 4-7 | 5-8 | 4-7 | 4.75-8 |
|  | Missing (n, %) | 20 (3.1) | 5 (4.1) | 0 | 7 (4.2) | 8 (2.5) | 0 |
| **Fatigue severity n (%)** | Clinically significant ($\geq 6/10$) | 333 (51.1) | 60 (48.8) | 7 (58.3) | 97 (58.4) | 147 (46.4) | 22 (64.7) |
| | Not clinically significant ($< 6/10$) | 299 (45.9) | 58 (47.2) | 5 (41.7) | 62 (37.3) | 162 (51.1) | 12 (35.3) |
|  | Missing | 20 (3.1) | 5 (4.1) | 0 | 7 (4.2) | 8 (2.5) | 0 |

**Figure 1.**
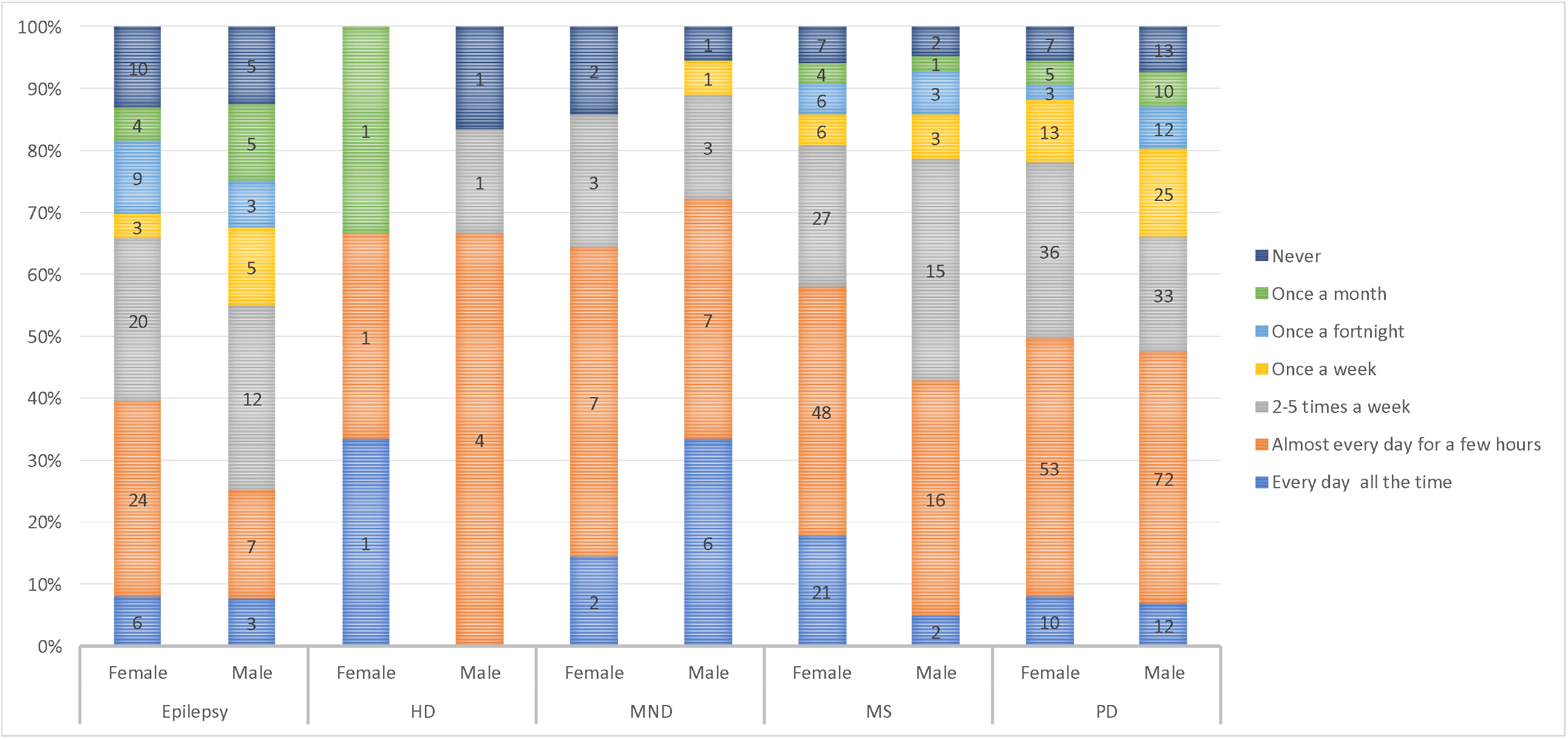
Reported frequency of fatigue (“How often do you experience fatigue?”). Percentages are normalised within each condition-sex group to show relative frequency distributions. MS n=166, PD n=317, Epilepsy n=123, MND n=34, HD n=12.

### Functional impact and help seeking

Fatigue frequently disrupted daily functioning. Overall, 38.5% reported social life impact and 34.5% concentration difficulties ‘often’ or more frequently (Table 2). The highest rates of social impact were observed in MND (58.7%), while MS (44.0%) and epilepsy (41.5%) were most often affected in terms of concentration.

Despite the high burden of fatigue, most participants had never discussed it with a clinician. Among respondents to the help-seeking question (n=603), 69.8% (n=421) reported no prior discussion, with particularly high non-disclosure in Parkinson’s disease (73.5%) and epilepsy (70.7%). Importantly, this gap persisted even among those with clinically significant fatigue: of participants reporting fatigue severity ≥6/10 who answered the help-seeking question (n=328), 60.4% had not sought professional advice, including over 70% of those with Parkinson’s disease and epilepsy. Together, these findings demonstrate a substantial disconnect between fatigue burden and clinical recognition across neurological conditions.

Multidimensional fatigue profiles across conditions, spanning severity, frequency, functional impact, and reporting, are illustrated in Figure 2.

**Figure 2.**
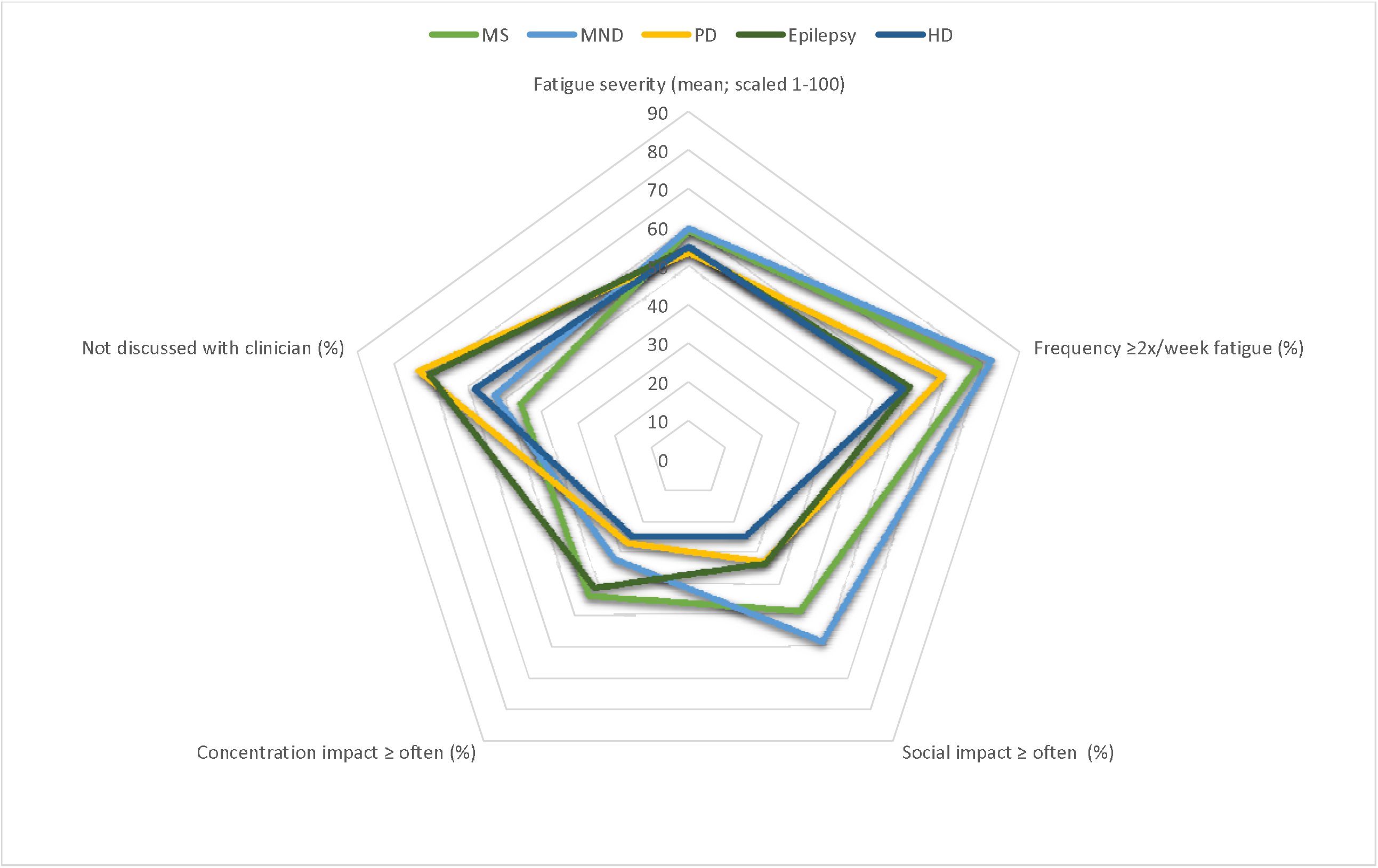
Fatigue impact profiles across neurological conditions. Fatigue severity (1–10 numerical rating scale) is rescaled to 0–100 for comparability with other dimensions. MS n=166, PD n=317, Epilepsy n=123, MND n=34, HD n=12.

### Fatigue severity

Self-rated fatigue severity was approximately normally distributed (mean 5.5, SD 2.2). Using a ≥6/10 threshold (aligned with MS/PD literature [40, 41]), 51.1% met the criteria for clinically significant fatigue. Severity was highest in MS and MND. HD also showed high mean scores, although estimates were imprecise due to the small sample, while epilepsy showed the widest dispersion of scores. In univariable analyses, history items (episode frequency; frequency of social and concentration impact) were strongly associated with severity, whereas sex, living status, and employment were not (Table 3).

**Table 3.** Regression outputs of the unadjusted and adjusted full specification linear model (complete cases n=481).

| Variable | Unadjusted model (n=652) |  |  | Adjusted model (n=481) |  |  |
| --- | --- | --- | --- | --- | --- | --- |
|  | Univariate R<sup>2</sup> statistic | Unstandardised B coefficient (95% CI) | p value | Unstandardised B coefficient (95% CI) | p value | Adjusted (BC 95%CI) |
| **Intercept** |  |  |  | 1.81 (1.15 to 2.46) | **<0.001** | 1.20 to 2.44 |
| **Condition** | 0.0138 |  |  |  |  |  |
| Epilepsy |  | 0.14 (-0.32 to 0.60) | 0.541 | -0.14 (-0.64 to 0.36) | 0.580 | -0.62 to 0.31 |
| Huntington's disease |  | 0.16 (-1.09 to 1.41) | 0.801 | -0.02 (-1.30 to 1.27) | 0.980 | -1.98 to 1.48 |
| Multiple sclerosis |  | 0.57 (0.16 to 0.99) | **0.007** | 0.21 (-0.22 to 0.64) | 0.343 | -0.29 to 0.64 |
| Motor neurone disease |  | 0.63 (-0.14 to 1.40) | 0.107 | 0.03 (-0.69 to 0.75) | 0.929 | -0.48 to 0.65 |
| Parkinson's disease |  | Reference |  | Reference |  | Reference |
| **Age** | 0.0217 |  |  |  |  |  |
| 18-34 |  | 1.22 (0.54 to 1.91) | **0.001** | 0.71 (-0.14 to 1.55) | 0.100 | -0.18 to 1.62 |
| 35-44 |  | 0.38 (-0.37 to 1.12) | 0.319 | 0.11 (-0.64 to 0.86) | 0.771 | -0.58 to 0.81 |
| 45-54 |  | 0.33 (-0.17 to 0.84) | 0.194 | -0.005 (-0.56 to 0.55) | 0.986 | -0.51 to 0.54 |
| 55-64 |  | 0.30 (-0.16 to 0.75) | 0.206 | 0.31 (-0.11 to 0.72) | 0.146 | -0.08 to 0.71 |
| ≥65 |  | Reference |  | Reference |  | Reference |
| **Gender** |  |  |  |  |  |  |
| Male | 0.0016 | -0.18 (-0.52 to 0.17) | 0.317 | 0.15 (-0.16 to 0.45) | 0.333 | -0.16 to 0.47 |
| Female |  | Reference |  | Reference |  | Reference |
| **Live alone** |  |  |  |  |  |  |
| No |  | Reference |  | Reference |  | Reference |
| Yes | 0.0007 | -0.14 (-0.57 to 0.29) | 0.522 | -0.22 (-0.58 to 0.14) | 0.238 | -0.58 to 0.13 |
| **Working status** |  |  |  |  |  |  |
| Working |  | Reference |  | Reference |  | Reference |
| Retired | 0.0001 | 0.05 (-0.32 to 0.42) | 0.795 | -0.05 (-0.49 to 0.38) | 0.808 | -0.47 to 0.38 |
| **Comorbidities** | 0.0145 |  |  |  |  |  |
| None |  | Reference |  | Reference |  | Reference |
| One |  | -0.01 (-0.49 to 0.32) | 0.692 | -0.08 (-0.42 to 0.27) | 0.654 | -0.43 to 0.24 |
| Two |  | 0.20 (-0.31 to 0.71) | 0.442 | -0.19 (-0.62 to 0.24) | 0.382 | -0.60 to 0.26 |
| ≥ 3 |  | 0.87 (0.26 to 1.49) | **0.006** | 0.07 (-0.45 to 0.59) | 0.797 | -0.35 to 0.51 |
| **Duration since diagnosis (years)** | 0.0002 | 0.003 (-0.01 to 0.019) | 0.753 | 0.02 (0.01 to 0.03) | **0.003** | 0.006 to 0.034 |
| **Frequency** | 0.4004 |  |  |  |  |  |
| Every day, all the time |  | 5.34 (4.70 to 5.98) | **<0.001** | 3.20 (2.21 to 4.19) | **<0.001** | 2.05 to 4.33 |
| Almost every day for a few hours |  | 4.03 (3.49 to 4.56) | **<0.001** | 2.35 (1.45 to 3.24) | **<0.001** | 1.31 to 3.31 |
| 2-5 times per week |  | 3.22 (2.66 to 3.78) | **<0.001** | 1.73 (0.84 to 2.63) | **<0.001** | 0.81 to 2.82 |
| Once a week |  | 2.48 (1.82 to 3.15) | **<0.001** | 1.28 (0.34 to 2.22) | **0.008** | 0.33 to 2.33 |
| Once a fortnight |  | 1.79 (1.04 to 2.53) | **<0.001** | 0.90 (-0.10 to 1.90) | 0.079 | -0.19 to 2.05 |
| Once a month |  | 1.05 (0.26 to 1.84) | **0.009** | 0.15 (-0.84 to 1.14) | 0.764 | -0.83 to 1.05 |
| Never |  | Reference |  | Reference |  | Reference |
| **Social life** | 0.3596 |  |  |  |  |  |
| All the time |  | 4.58 (3.96 to 5.21) | **<0.001** | 1.89 (1.02 to 2.77) | **<0.001** | 0.93 to 2.89 |
| Most of the time |  | 3.83 (3.25 to 4.41) | **<0.001** | 1.50 (0.68 to 2.32) | **<0.001** | 0.61 to 2.41 |
| Often |  | 3.30 (2.75 to 3.85) | **<0.001** | 1.33 (0.55 to 2.10) | **0.001** | 0.45 to 2.15 |
| Sometimes |  | 2.61 (2.11 to 3.12) | **<0.001** | 0.99 (0.28 to 1.71) | **0.006** | 0.03 to 1.71 |
| Rarely |  | 1.19 (0.61 to 1.75) | **<0.001** | 0.24 (-0.47 to 0.95) | 0.509 | -0.57 to 0.98 |
| Never |  | Reference |  | Reference |  | Reference |
| **Concentration** | 0.2684 |  |  |  |  |  |
| All the time |  | 4.02 (3.34 to 4.71) | **<0.001** | 1.34 (1.01 to 2.49) | **0.002** | 0.29 to 2.35 |
| Most of the time |  | 3.88 (3.24 to 4.52) | **<0.001** | 1.07 (0.32 to 1.82) | **0.005** | 0.33 to 1.98 |
| Often |  | 2.91 (2.34 to 3.49) | **<0.001** | 0.71 (0.04 to 1.37) | **0.037** | -0.21 to 1.58 |
| Sometimes |  | 2.23 (1.71 to 2.76) | **<0.001** | 0.62 (0.01 to 1.23) | **0.047** | -0.16 to 1.43 |
| Rarely |  | 1.56 (1.00 to 2.13) | **<0.001** | 0.24 (-0.38 to 0.86) | 0.453 | -0.48 to 0.97 |
| Never |  | Reference |  | Reference |  | Reference |

### Multivariable model

#### Full model

In multivariable analysis (complete cases, n = 481), three history items, (i) episode frequency, (ii) frequency of social-life impact (iii) frequency of concentration impact, were independent predictors of severity after adjusting for demographics, diagnosis, comorbidity, living/working status, and duration since diagnosis. Episode frequency had the largest effect size. Demographic, diagnostic, and contextual covariates showed small, non-significant effects, except for duration since diagnosis which showed a modest positive association.

Variable selection was based on complete-case analysis to ensure transparent identification of the most informative predictors. The final model was then re-estimated using multiple imputation (m = 20; n=652) to assess robustness to missing data. Imputed and complete-case models yielded highly consistent coefficients and significance patterns, confirming model stability (see Supplementary files).

In the apparent sample (n = 481), the full model explained 49.18% of variance in fatigue severity (adjusted R^2^ = 0.4918; RMSE = 1.53; MAE = 1.19). Calibration was assessed using 10-fold cross-validation and showed close agreement between predicted and observed scores (adjusted R^2^ = 0.45; RMSE = 1.59; MAE = 1.27; slope = 0.92, BC 95% CI 0.84 to 1.01; intercept +0.39, BC 95% CI −0.06 to +0.91) indicating good overall calibration (Figure 3).

**Figure 3.**
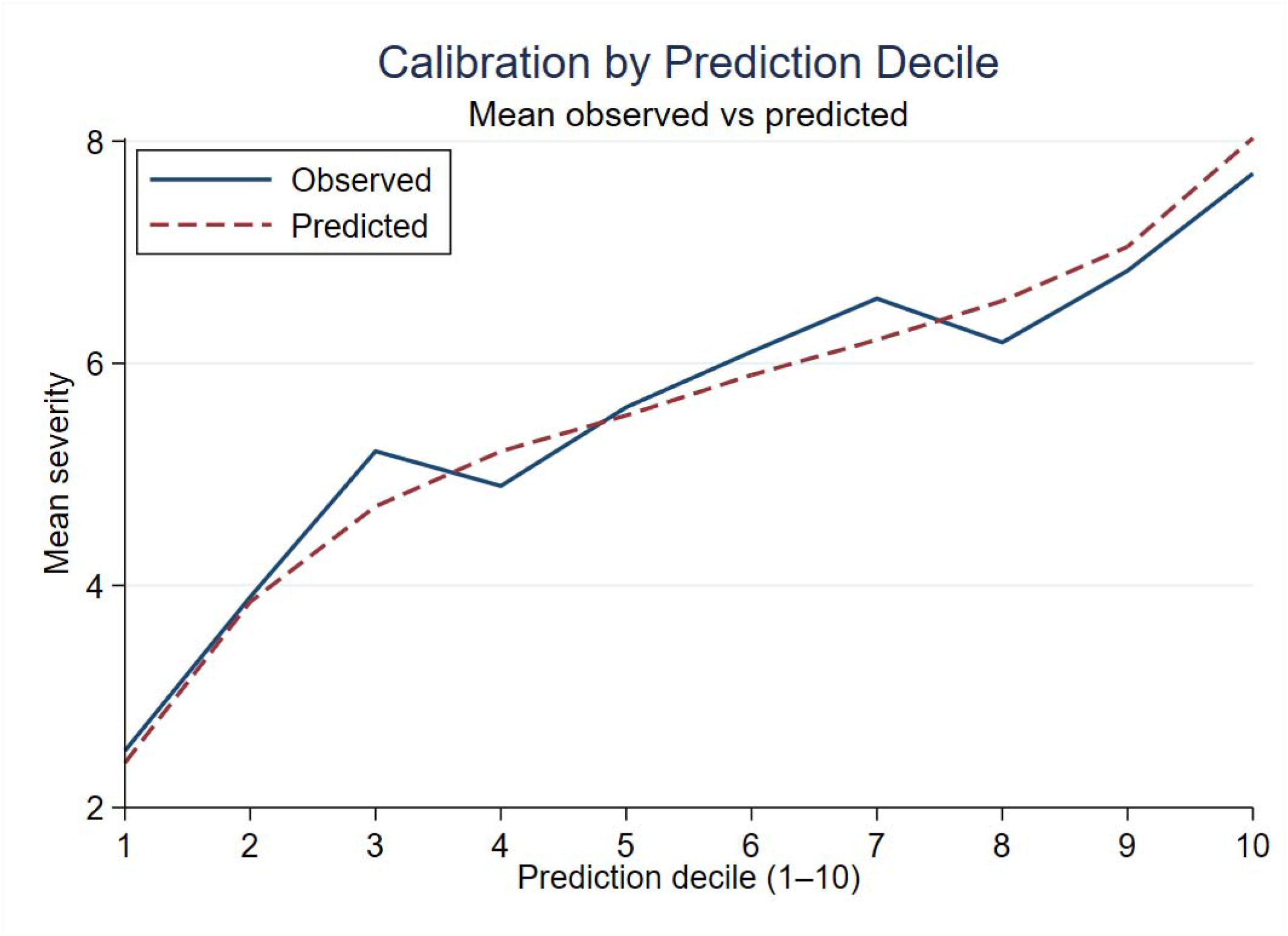
Calibration of the full multivariable model using 10-fold cross-validation (predicted versus observed fatigue severity).

#### Three-item model

As most predictive signal resided in the three history items, we specified a simplified model restricted to these variables; regression coefficients for each response category are shown in Table 4. Using the same complete-case dataset (*n* = 481), performance remained comparable to the full model; apparent sample adjusted R^2^ = 0.49; RMSE = 1.53; MAE = 1.21, confirming that brief self-reported fatigue history captured nearly all the predictive information present in the full model. Cross-validated performance was similar (adjusted R^2^ = 0.47; RMSE = 1.57; MAE = 1.25). Calibration remained good (slope = 0.96, BC 95% CI 0.87–1.05; intercept = +0.21, −0.30 to +0.73), indicating good agreement between predicted and observed values, and minimal loss of accuracy compared with the full model (Figure 4).

**Table 4.** Coefficients for the three-item fatigue history model (complete cases n=481)

| Item | Response categories | $\beta$ coefficient |
| --- | --- | --- |
| 1. How often have you felt fatigued in the past month? | Never | Reference |
|  | Once a month | +0.21 |
|  | Once a fortnight | +0.85 |
|  | Once a week | +1.29 |
|  | 2-5 times per week | +1.70 |
|  | Almost every day for a few hours | +2.25 |
|  | Every day, all the time | +3.16 |
| 2. Does fatigue affect your social life? | Never | Reference |
|  | Rarely | +0.16 |
|  | Sometimes | +0.90 |
|  | Often | +1.17 |
|  | Most of the time | +1.36 |
|  | All the time | +1.64 |
| 3. Does fatigue affect your concentration? | Never | Reference |
|  | Rarely | +0.32 |
|  | Sometimes | +0.70 |
|  | Often | +0.82 |
|  | Most of the time | +1.16 |
|  | All the time | +1.54 |
|  | Constant (intercept) | 2.19 |

**Clinical case:** A young adult with epilepsy reports fatigue twice a week, sometimes affecting social life and most of the time affecting concentration.

**Figure 4.**
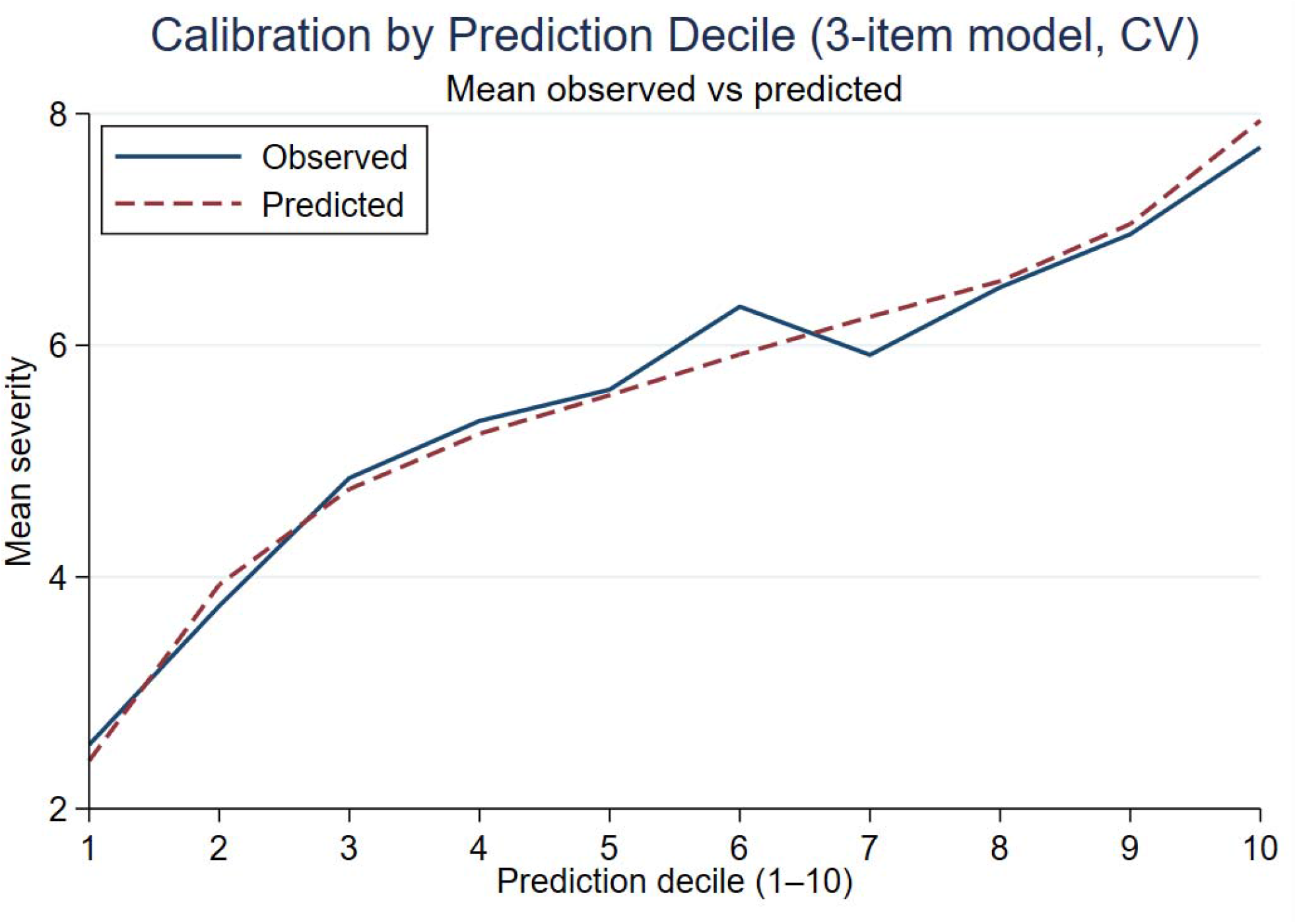
Calibration of the three-item history-based model using 10-fold cross-validation (predicted versus observed fatigue severity).

To assess robustness, the same three-item model was also tested in all participants with complete data on the three history items and fatigue severity (n = 581). Results were highly consistent (CV RMSE= 1.58; MAE = 1.26; slope = 0.97, BC 95% CI 0.89–1.04; intercept = +0.20, −0.27 to +0.66), confirming that predictive accuracy and calibration was consistent across samples. Taken together, these findings suggest that the three history questions capture the dominant drivers of fatigue severity, and that model performance is robust to missingness in contextual variables.

For clinical interpretability, predicted scores were grouped into three pragmatic severity bands:

- **Low (<4):** infrequent or low-impact fatigue
- **Moderate (4.0–5.9):** intermittent fatigue with some disruption
- **High (**≥**6):** frequent, functionally disruptive fatigue

Example clinical vignette:

At annual review, a young adult with epilepsy reports fatigue episodes twice a week, sometimes affecting social life and most of the time affecting concentration. Applying the three-item model yields a predicted score of 5.96 (Table 4), placing the patient in the high-severity band (≥6). This illustrates how a brief, structured history can reveal clinically important fatigue that might otherwise remain unrecognised in routine care.

## Discussion

This study has two main findings. Using a single measurement approach across five neurological conditions, we quantified fatigue severity, episode frequency and the frequency with which fatigue disrupts social participation and concentration, dimensions often hidden by heterogeneous tools and composite scores. Second, we derived a three-item model that explains almost half the variance in self-rated severity with good internal calibration and performance similar to the full model. Together, these findings advance understanding of fatigue across conditions and support a shift from descriptive fatigue scales toward brief, transdiagnostic indicators suitable for both clinical and digital use.

To our knowledge, this is the first study to compare severity, episode frequency, and functional impact of fatigue across five neurological disorders using one instrument. Fatigue was common and functionally disruptive in all groups, with particularly heavy burden in people with MND and MS. About two-thirds of participants had never discussed fatigue with a clinician, highlighting the enduring disconnect between lived burden and clinical recognition. Through structured item reduction, three questions (episode frequency, and the frequency of impact on social life and concentration) emerged as the most consistent predictors across diagnoses and appear to index a latent construct of fatigue severity. Pending external validation and threshold optimisation, this brief history-based tool could be incorporated into routine and digital pathways to streamline triage and support earlier, more personalised care in neurology and potentially other long-term conditions.

### Comparative fatigue severity and episode frequency across conditions

#### Prevalence and diagnostic patterns

The prevalence of clinically significant fatigue aligned with previous literature across chronic conditions [3–7, 12, 45–49]. It was highest in MND, followed by MS and HD, and lower in epilepsy and PD, broadly consistent with a large prospective cohort study (n=78,363) reporting severe fatigue in 54% of people with MS (n=156), 40% in PD (n=78), and 31% in epilepsy (n=940) [50]. Our estimate for HD was lower than the only comparable report to date (82.5%, n=165) [51], which may reflect our smaller HD sample (n=12).

Prevalence was higher in epilepsy (48.8%; n=123), consistent with a recent systematic review [4]. The only other comparative study, to our knowledge, using the same scale reported similar mean fatigue scores for epilepsy and MS, although the primary outcome was to assess different rating scales and sample sizes were small [52].

#### Frequency of fatigue episodes: a neglected dimension

Fatigue was not only prevalent, but episodes were also frequent. Nearly half of participants experienced fatigue lasting ‘almost every day for a few hours’ or more often. Frequency was highest in MND (82.3% reporting ≥2 episodes per week) and lowest in epilepsy (35.8% reporting episodes of fatigue once per week or less). In PD (n=317), previous studies found that 67% experienced fatigue twice a week or more often, comparable to the 69.2% reported here [16]. For MS (n=166), our findings matched previous smaller studies in the literature [53] [54].

Frequency of fatigue episodes has not been reported to our knowledge in epilepsy, HD or MND and there have been no previous studies comparing episode frequency across five different neurological conditions using the same instrument. This is an important gap, as frequency may capture clinically meaningful variation that severity or global scores alone can obscure. Despite its potential relevance for monitoring, prognosis, and treatment planning, episode frequency is rarely included in widely used fatigue measures and currently lacks standardised clinical thresholds. Earlier mixed-methods work in Parkinson’s disease first highlighted that higher fatigue frequency was associated with greater disruption to daily activities, social participation and concentration, and that fatigue was rarely discussed with clinicians [55, 56].

### Functional impact in social and cognitive domains

#### Social participation

Across the cohort, 38.5% of participants reported fatigue affecting their social life ‘often’ or more frequently. This proportion is consistent with, but adds a temporal dimension to, earlier work that linked higher fatigue scores to poorer social participation,[25–28, 55, 56]. Social burden was greatest in MND likely reflecting the compounded effects of fatigue and disease progression, whereas people living with epilepsy most often selected ‘never’, suggesting that episodic fatigue in this condition may be less socially limiting or perhaps that its effects are overshadowed by other seizure-related restrictions.

#### Concentration

Fatigue also compromised concentration, with about a third of the total sample reporting impact ‘often’ or more frequently. Despite differing overall profiles, MS and epilepsy showed similarly high rates of frequent concentration impact (44% and 41.5% respectively).

In MS, this is consistent with several studies reporting a negative correlation between fatigue and cognitive performance [58–59] and an MS digital app (n=2095) which reported that the most common concern was the cluster of fatigue and difficulties with concentration [31].

In epilepsy, the pattern was particularly striking. Participants reported the lowest frequency of fatigue episodes yet similar levels of concentration impact to MS. This suggests that even intermittent fatigue may be perceived as cognitively disruptive in epilepsy, especially among younger, working-age adults. Potential contributors include peri-ictal fatigue, antiepileptic drug effects, sleep disturbance, or underlying cognitive vulnerabilities. As our data are self-reported, we cannot confirm that concentration problems were solely attributable to fatigue and further work is needed to clarify mechanisms and temporal relationships.

### Distinct phenotypic profiles by condition

Distinct patterns of fatigue impact emerged across conditions. In MND, social life appeared more affected than concentration, whereas in epilepsy the pattern was reversed. In HD, despite relatively high self-rated severity, fewer participants reported frequent functional impact, which may reflect reduced symptom awareness, although interpretation is limited by the small sample and higher missingness. In PD, concentration impact was lowest despite almost half reporting daily fatigue, while social impact was comparable to epilepsy, possibly reflecting coping, normalisation in older adults, or under-recognition. MS showed the broadest burden, with high social and cognitive impact alongside frequent daily fatigue, highlighting fatigue in MS as a pervasive, multidomain challenge. Taken together, these profiles highlight opportunities to personalise fatigue support, as different patterns of social and cognitive impact may influence help-seeking and preferences for assistance.

### Help-seeking and the hidden burden

Despite the high prevalence of clinically significant fatigue across the whole sample, 69.8% of respondents to the help-seeking question (421/603) had never discussed their fatigue with a clinician. Non-disclosure was particularly common in PD, as has been demonstrated previously [55, 56], and epilepsy, but was also substantial in MS, where fewer than half of participants had sought clinical advice despite reporting the highest combined levels of fatigue prevalence, frequency and functional impact. Overall, around three in five participants with clinically significant fatigue had not sought professional advice, indicating a substantial gap between symptom burden and clinical recognition.

This pattern aligns with previous findings of fatigue underreporting in neurological conditions [60] and likely reflects a combination of patient-related barriers (e.g. uncertainty about whether fatigue is a treatable symptom or whether it is related to their condition), clinical challenges (e.g. limited tools and time to assess fatigue, perception of limited treatment options), and wider system-level factors.

Regardless of cause, these data highlight a significant unmet clinical need and represent a missed opportunity for early intervention to prevent functional decline, improve quality of life and reduce potentially avoidable downstream health service utilisation. They also underscore the need for brief, pragmatic tools that normalise discussion of fatigue in time-limited consultations.

### Estimating fatigue severity: a pragmatic approach

Given the high levels of under-reporting and functional disruption, we aimed to derive a simple, functionally grounded model to support earlier recognition of fatigue. From a broad set of potential predictors, including demographics, diagnosis, comorbidities and multiple fatigue related items, our regression analysis identified three self-reported items as the strongest independent predictors of fatigue severity: frequency of fatigue episodes, plus how often fatigue impacts social life and concentration. Together, these explained nearly half of the variance in fatigue severity with good predictive accuracy and minimal overfitting. The close correspondence between the full and three-item models indicates that brief history-taking captures most of the predictive information relevant to perceived fatigue severity. These three items appear to index a latent construct of fatigue severity in our dataset, providing a concise psychometric core that is both clinically intuitive and potentially transportable across conditions.

The three-item model has several advantages. Traditional fatigue assessments tend to rely on Likert-style rating scales, requiring individuals to quantify how strongly they agree with statements such as ‘Fatigue interferes with my physical functioning.’ These formats can be cognitively demanding, particularly for people with neurological conditions, and may not reflect how fatigue is experienced or communicated in daily life.

By contrast, our model draws on simple, concrete questions about how often episodes of fatigue occur and how frequently they affect concentration and social life, domains that are familiar, observable, and clearly relevant to patients. Frequency is a particularly accessible construct; it is often easier to recall how often something happens than to assign a number or judge level of agreement with an abstract statement. These questions can be incorporated naturally into conversation during routine consultations without feeling like a formal test. This approach may support more consistent responses, improve patient engagement, and enhance the clinical utility of fatigue assessment in specialist and community settings, both in-person and remotely.

Episode frequency was the strongest predictor. Yet most fatigue scales (e.g., PROMIS Fatigue, FACIT-F [61]) rarely capture how often fatigue occurs or how often it disrupts daily activities. As the construct validity of some widely used fatigue scales has recently been questioned [62, 63], there may be value in complementing established measures with brief, patient-derived indicators that capture episode frequency and tangible, real-world disruption.

Social and cognitive impact made independent contributions to perceived fatigue severity, indicating that they are not simply proxies of fatigue frequency but are weighted separately by patients. There was a clear dose-response, with higher frequencies showing the largest effects. Their explanatory importance is supported by recent machine learning work in PD, which identified ‘fatigue is among my three most troubling symptoms’ and ‘fatigue interferes with my work, family or social life’ as key predictors from the nine items on the Fatigue Severity Scale [64].

Most demographics, comorbidities, and work status were not predictive in our adjusted model, echoing inconsistent associations across the literature [3, 6, 7, 45, 47, 52, 66–67]. Notably, neurological diagnosis was also not a significant predictor. This is consistent with the growing recognition that fatigue may be underpinned by shared transdiagnostic mechanisms which operate across a broad range of chronic diseases, not only neurological disorders (e.g. network dysfunction, inflammation, metabolic disturbance) [50, 68, 69]. Taken together, these results reinforce the case for moving beyond diagnosis-led approaches to fatigue assessment and instead focusing on symptom burden and functional impact.

### Clinical implications

Fatigue contributes to poorer treatment adherence, loss of independence, and escalating health needs, yet it remains under-assessed in routine neurological care [37, 70]. In the UK, neurology admissions are rising faster than in any other specialty. The average cost of a planned neurological admission in Wessex is £1537 [71], with unplanned admissions costing the NHS over £120 million annually [37]. Earlier recognition and more proactive management of fatigue may help reduce avoidable crisis points [36] and reduce downstream system pressure.

Our findings highlight the high prevalence and functional burden of fatigue across neurological conditions, with distinct patterns by diagnosis. In our data, fatigue impact emerged as predominantly cognitive in epilepsy, socially disruptive in MND, and pervasive across domains in MS. Recognising this phenotypic heterogeneity may support more tailored, person-centred management strategies that improve quality of life, social participation, and functional independence. Routine screening may also enhance treatment adherence and clinic attendance, both vital for improving outcomes [72],[73].

Importantly, almost all the predictive information contained in the full multivariable model was captured by just three history questions about fatigue frequency and frequency of impact on social life and concentration. The three-item model offers clinicians an intuitive triage mechanism that can be completed in under a minute. It could be useful for routine reviews, could be embedded in digital care pathways (patient portals, diaries for pre-clinic assessments), and could potentially be adaptable to other long-term conditions such as long COVID [69, 74], cancer-related fatigue [75], and autoimmune diseases [68]. By providing a consistent, brief metric across conditions, this model could support more standardised fatigue assessment in both clinical practice and research, analogous to the role of very short mental health screeners (e.g. PHQ-2, GAD-2). The full regression model may be valuable for specialist use and clinical trials. In line with TRIPOD guidance, however, external validation in external datasets will be the essential next step to assess calibration, optimise cut points, and confirm generalisability.

## Limitations

This cross-sectional survey permits only associative inference; longitudinal studies are required to examine trajectories and prognostic value. Convenience sampling via clinics and condition-specific support groups may have over-represented motivated individuals and under-represented those with more advanced disease or disengagement from services. All variables were self-reported, with potential recall and misclassification error. However, our prevalence and severity patterns align closely with external literature, supporting external validity.

To minimise participant burden, we did not measure additional correlates of fatigue such as depression, sleep disturbance, pain, or medication class. These factors are known to overlap with fatigue both conceptually and statistically, so residual confounding cannot be excluded. However, our aim was not to disentangle aetiological mechanisms but to quantify burden consistently across conditions and develop a brief triage tool. In this context, overlap with related constructs reflects lived experience and may strengthen the clinical utility of the model.

The relatively small HD and MND samples reduce the precision of subgroup estimates, although this partly reflects the rarity of these conditions. In addition, our model was developed and banded within a single dataset. External validation is necessary and at present, applicability to other long-term conditions remains hypothesised but unproven.

## Conclusions

Fatigue is common, burdensome, and persistently under-recognised across neurological conditions. We describe a brief, patient-derived model comprising three history questions that quantify how often fatigue occurs and how often it interferes with social life and concentration. Pending external validation and refinement of thresholds, this approach offers a pragmatic, low-burden alternative to lengthy scales and a clinically meaningful prompt for more detailed assessment. It may support earlier recognition, more personalised management, and improved quality of life for people living with neurological and other long-term conditions in which fatigue is a major challenge. As global efforts increasingly prioritise patient-reported outcomes and digital triage, a simple history-based model that captures the latent construct of fatigue severity may provide a scalable foundation for harmonising fatigue assessment across research and clinical contexts. Future work should focus on external validation in independent datasets, including both neurology and non-neurological long-term conditions, testing sensitivity to change in intervention studies, and integrating this approach into digital health systems to monitor fatigue trajectories over time.

## Data Availability

The datasets generated and/or analysed during the current study are available from the corresponding author on reasonable request.

## Abbreviations

BC: Bias-corrected
CI: Confidence interval
CV: Cross-validation / Cross-validated
df: Degrees of freedom
ERGO: Ethics and Research Governance Online (University of Southampton)
FACIT: Functional Assessment of Chronic Illness Therapy (Fatigue)
FSS: Fatigue Severity Scale
HD: Huntington’s disease
IRAS: Integrated Research Application System (UK)
IQR: Interquartile range
MAE: Mean absolute error
MFIS: Modified Fatigue Impact Scale
MND: Motor neurone disease
MS: Multiple sclerosis
NRS: Numerical Rating Scale
OR: Odds ratio
PD: Parkinson’s disease
PROMIS: Patient-Reported Outcomes Measurement Information System
RMSE: Root mean squared error
R^2^: Coefficient of determination
SD: Standard deviation
SE: Standard error
U-FIS: Unidimensional Fatigue Impact Scale

## Declarations

### Ethics approval and consent to participate

Ethical approval for this study was granted by the NHS South Central Hampshire A Research Ethics Committee (IRAS 203783) and the University of Southampton Faculty of Medicine Ethics Committee (ERGO 57456). All participants gave informed consent prior to participation in the study.

### Consent for publication

Not applicable.

### Availability of data and materials

The datasets are available from the corresponding author on reasonable request. This work builds on a preliminary model developed by the lead author during MSc Public Health research (Fuller, 2023).

### CRediT authorship contribution statement

PF: Conceptualisation, methodology, data curation, formal analysis, visualisation, writing-original draft, writing – review and editing

SF: Investigation, writing – review and editing

BB: Conceptualisation, formal analysis, validation, writing – review and editing

SBP: Methodology, investigation, writing – review and editing

VA: Methodology, writing – review and editing

CK: Conceptualisation, methodology, funding acquisition, writing – review and editing.

All authors read and approved the final manuscript.

## Acknowledgements

We remain immensely grateful to all our participants who gave up their time and insight to participate in this survey to advance our understanding of fatigue, and to the support groups and NHS clinics who helped share the study with their networks.

---

**Supplementary Table S1.**
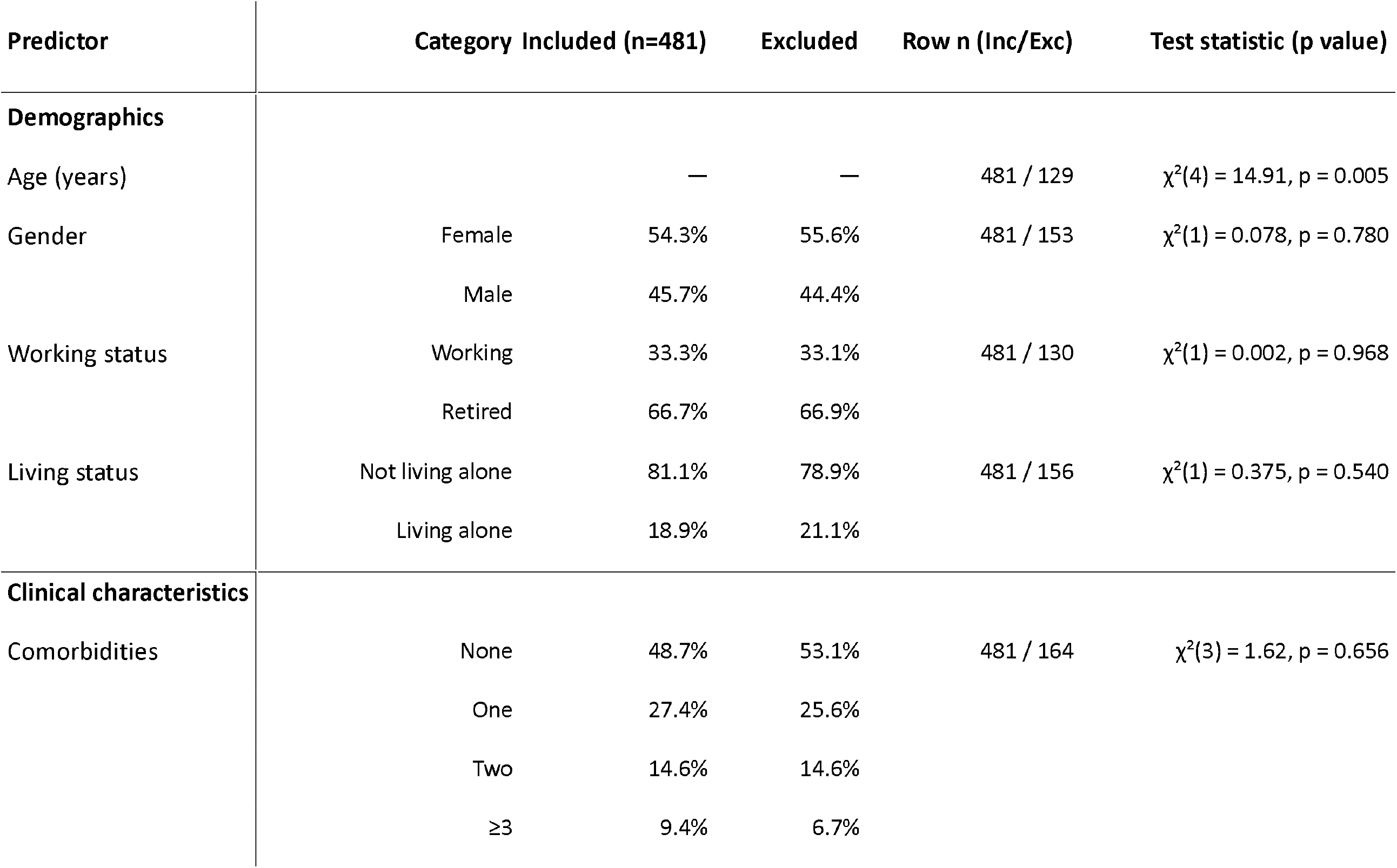
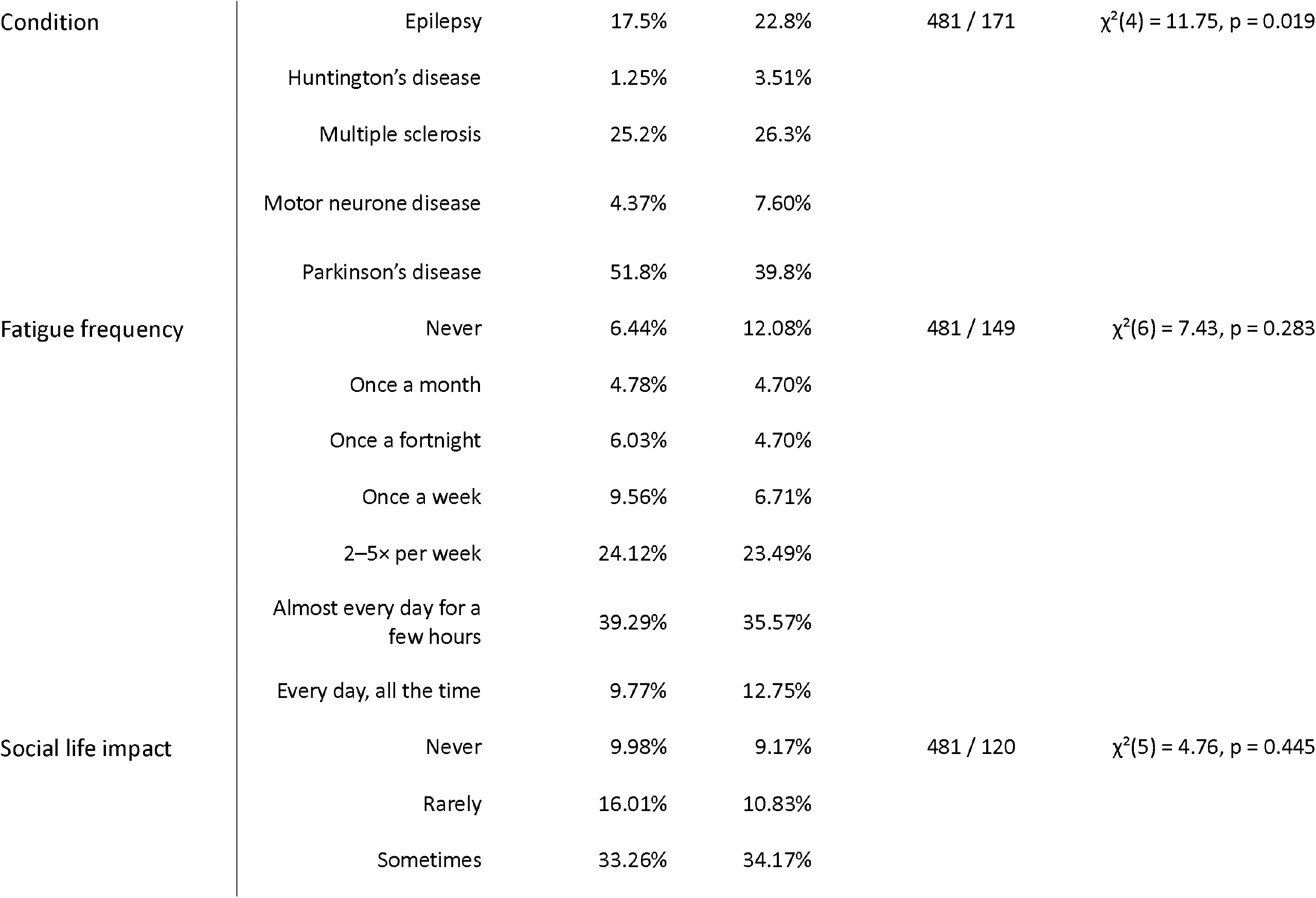
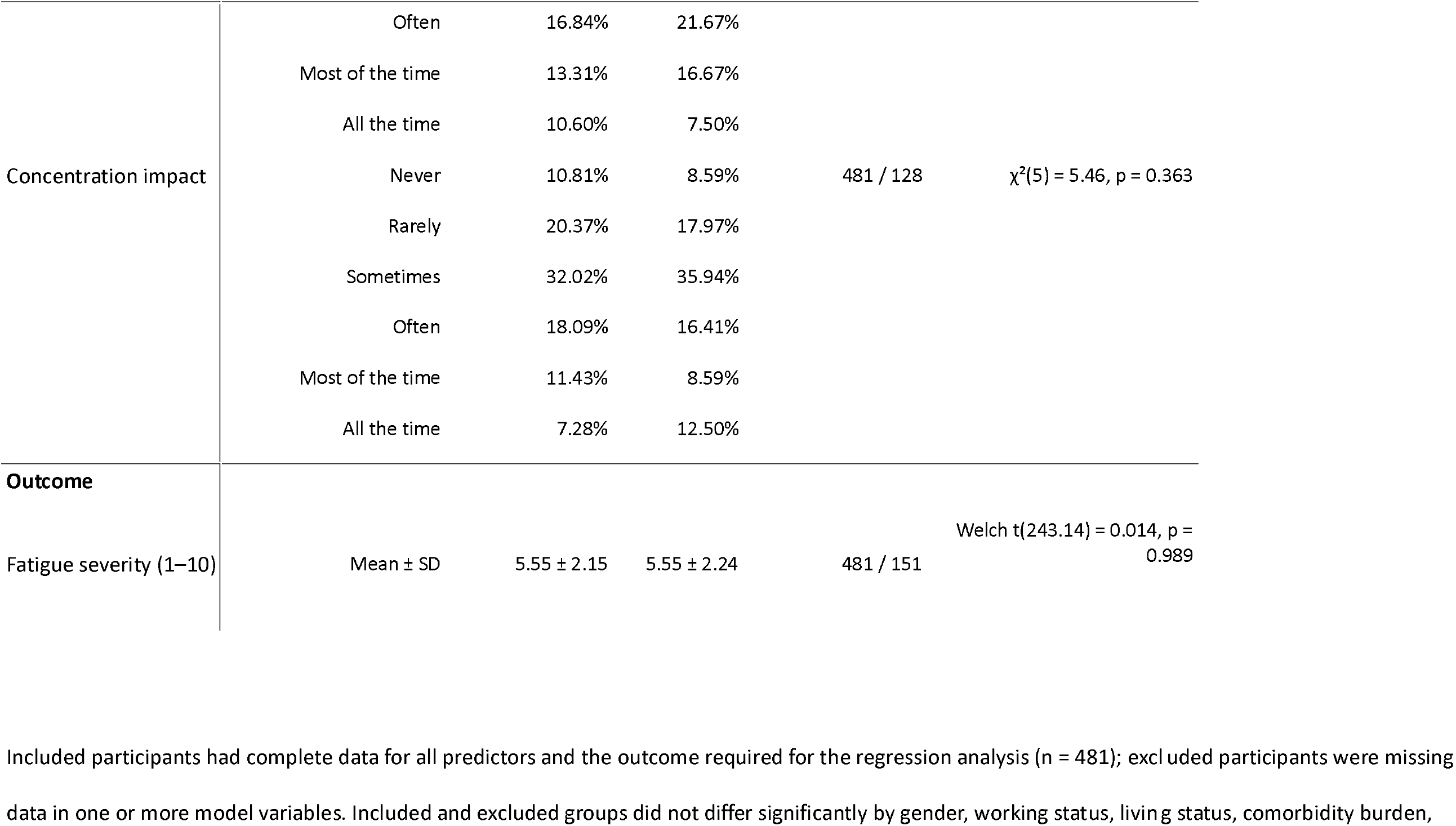
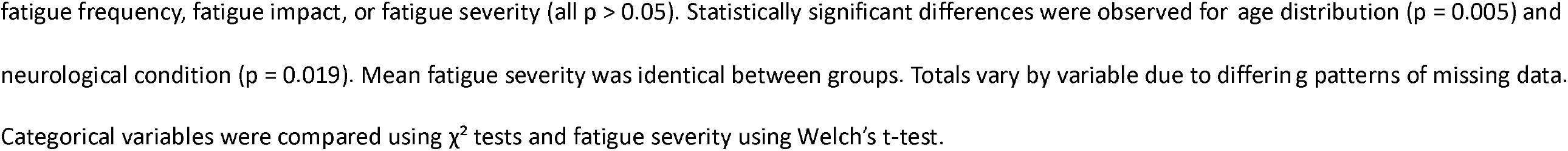
Comparison between complete-case (included) and excluded participants for the regression model.

**Supplementary Table S2:** Ordinal logistic regression for predictors of self-rated severity (1-10).

| Predictor | Category | Odds Ratio | 95% CI | p-value |
| --- | --- | --- | --- | --- |
| Fatigue frequency | Once a month | 2.69 | 0.59–12.35 | 0.203 |
|  | Once a fortnight | 4.51 | 0.86–23.51 | 0.074 |
|  | Once a week | 9.16 | 1.87–44.85 | 0.006 |
|  | 2–5 times a week | 15.08 | 3.12–72.94 | 0.001 |
|  | Almost every day for a few hours | 30.15 | 5.84–155.53 | <0.001 |
|  | Every day, all the time | 110.56 | 18.07–676.44 | <0.001 |
| Social impact | Rarely | 1.48 | 0.52–4.22 | 0.467 |
|  | Sometimes | 3.26 | 1.15–9.23 | 0.027 |
|  | Often | 4.71 | 1.51–14.64 | 0.008 |
|  | Most of the time | 5.70 | 1.70–19.15 | 0.005 |
|  | All the time | 10.55 | 2.63–42.34 | 0.001 |
| Concentration impact | Rarely | 1.46 | 0.50–4.26 | 0.489 |
|  | Sometimes | 2.02 | 0.68–5.97 | 0.205 |
|  | Often | 2.22 | 0.69–7.16 | 0.182 |
|  | Most of the time | 3.58 | 1.08–11.87 | 0.037 |
|  | All the time | 5.29 | 1.21–23.01 | 0.026 |
| Duration since diagnosis (years) | Continuous | 1.02 | 1.01–1.04 | 0.003 |
| Covariates | Age, gender, living alone, work, comorbidities, condition | All non significant | – | – |

Odds ratios (OR) and 95% confidence intervals (CI) are shown relative to the reference category for each variable. The model adjusted for age, gender, living status, employment, comorbidities, and neurological condition. Model summary: $n=481$ ; LR $\chi^2(31) = 356.09$ , $p < 0.001$ ; Pseudo $R^2 = 0.175$ ; AIC = 1762.2; BIC = 1929.2.

**Supplementary Table 3:**
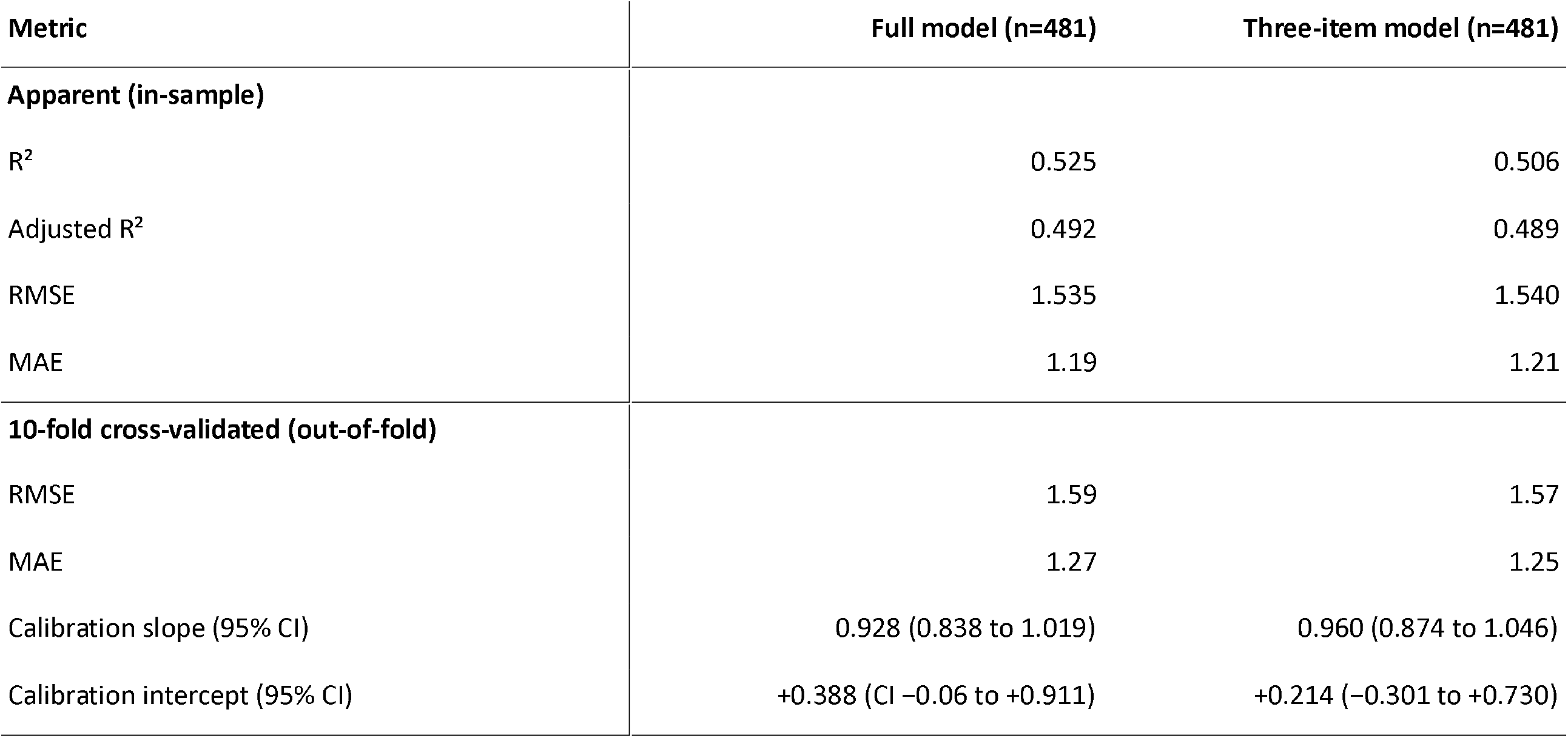
Model performance and calibration for full model and three-item model (complete cases n=481)

| Metric | Full model (n=481) | Three-item model (n=481) |
| --- | --- | --- |
| **Apparent (in-sample)** |  |  |
| R<sup>2</sup> | 0.525 | 0.506 |
| Adjusted R<sup>2</sup> | 0.492 | 0.489 |
| RMSE | 1.535 | 1.540 |
| MAE | 1.19 | 1.21 |
| **10-fold cross-validated (out-of-fold)** |  |  |
| RMSE | 1.59 | 1.57 |
| MAE | 1.27 | 1.25 |
| Calibration slope (95% CI) | 0.928 (0.838 to 1.019) | 0.960 (0.874 to 1.046) |
| Calibration intercept (95% CI) | +0.388 (CI -0.06 to +0.911) | +0.214 (-0.301 to +0.730) |

**Supplementary Table S4:** Multiple imputation sensitivity analysis (n = 652, m = 20).

| Predictor | Category | Beta (pooled) | 95% CI | p |
| --- | --- | --- | --- | --- |
| **Intercept** | – | 1.813 | 1.172 to 2.455 | <0.001 |
| **Gender** | Male | 0.212 | –0.054 to 0.477 | 0.118 |
| **Working status** | Retired | 0.054 | –0.362 to 0.470 | 0.800 |
| **Living status** | Yes (alone) | –0.312 | –0.633 to 0.010 | 0.058 |
| **Comorbidities** | One | –0.119 | –0.426 to 0.187 | 0.444 |
|  | Two | –0.282 | –0.662 to 0.098 | 0.146 |
|  | ≥3 | 0.247 | –0.219 to 0.712 | 0.299 |
| **Age** | 18–34 | 0.699 | –0.039 to 1.437 | 0.063 |
|  | 35–44 | 0.012 | –0.650 to 0.674 | 0.972 |
|  | 45–54 | –0.086 | –0.580 to 0.408 | 0.733 |
|  | 55–64 | 0.261 | –0.118 to 0.639 | 0.176 |
| **Condition** | Epilepsy | 0.029 | –0.421 to 0.479 | 0.899 |
|  | Huntington’s disease | 0.019 | –0.988 to 1.027 | 0.970 |
|  | Multiple sclerosis | 0.192 | –0.188 to 0.572 | 0.322 |
|  | Motor neurone disease | 0.014 | –0.575 to 0.603 | 0.963 |
| **Duration since diagnosis (years)** | per year | **0.015** | **0.002 to 0.027** | **0.022** |
| Frequency | Once a month | 0.212 | −0.667 to 1.091 | 0.636 |
|  | Once a fortnight | 0.568 | −0.329 to 1.465 | 0.214 |
|  | Once a week | **1.277** | **0.440 to 2.114** | **0.003** |
|  | 2–5× per week | **1.555** | **0.773 to 2.337** | **<0.001** |
|  | Almost every day for a few hours | **2.163** | **1.389 to 2.937** | **<0.001** |
|  | Every day, all the time | **2.968** | **2.112 to 3.824** | **<0.001** |
| Social life | Rarely | 0.312 | −0.330 to 0.954 | 0.341 |
|  | Sometimes | **1.172** | **0.551 to 1.794** | **<0.001** |
|  | Often | **1.486** | **0.816 to 2.157** | **<0.001** |
|  | Most of the time | **1.656** | **0.949 to 2.363** | **<0.001** |
|  | All the time | **2.102** | **1.337 to 2.866** | **<0.001** |
| Concentration | Rarely | 0.271 | −0.283 to 0.826 | 0.337 |
|  | Sometimes | **0.632** | **0.082 to 1.182** | **0.025** |
|  | Often | **0.785** | **0.174 to 1.397** | **0.012** |
|  | Most of the time | **1.164** | **0.478 to 1.850** | **0.001** |
|  | All the time | **1.159** | **0.433 to 1.884** | **0.002** |

Fatigue episode frequency, and the frequency with which fatigue affected social life and concentration, remained strong independent predictors of fatigue severity.

## Questionnaire used for all five neurological conditions

### Please answer as many questions as possible

#### SECTION 1: [Neurological Condition] and You

Please circle one answer for each question or fill in the box provided

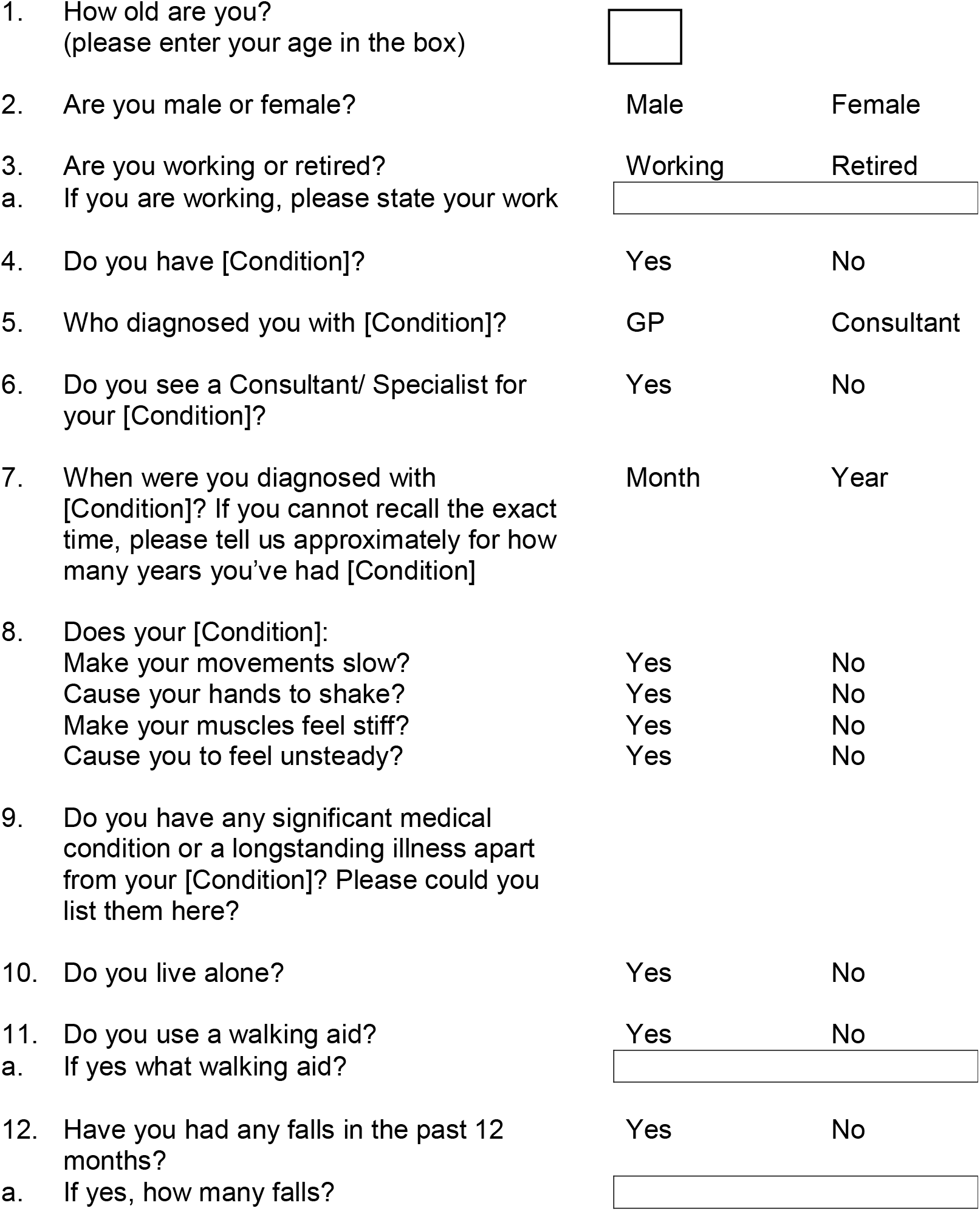
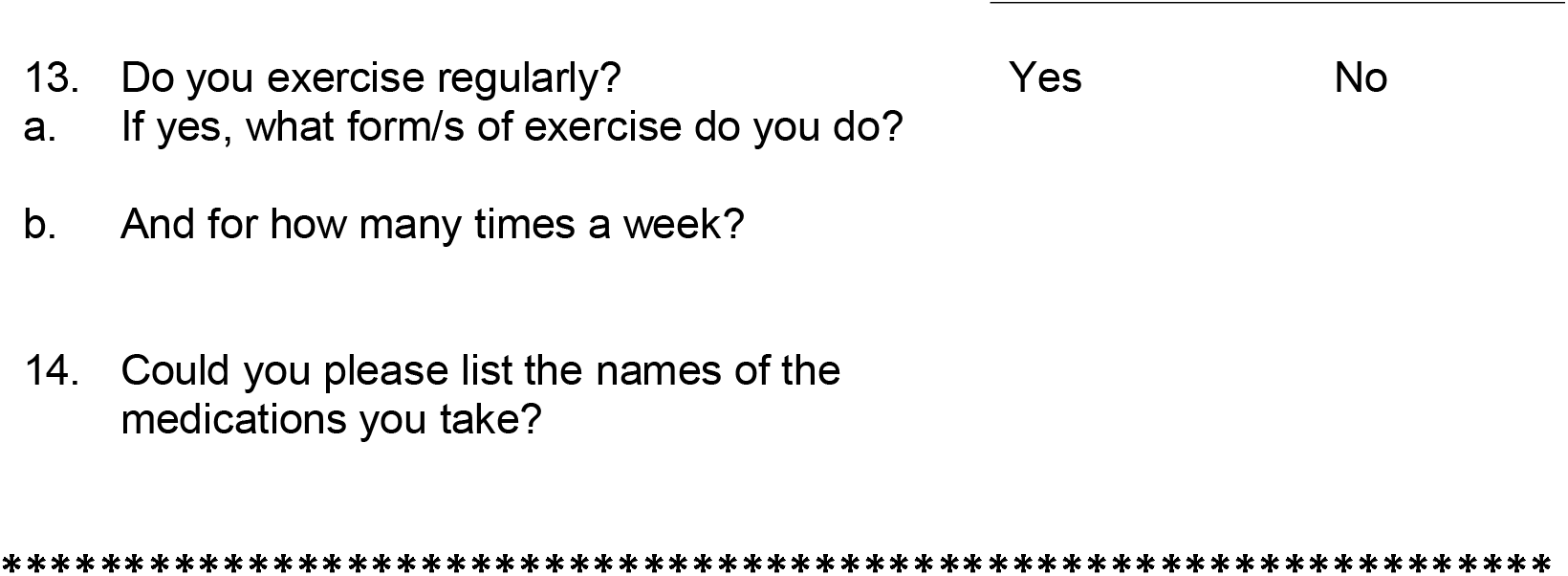

#### SECTION 2: Fatigue and how you describe it

The next few questions look at Fatigue. By this we mean intense or severe tiredness, or exhaustion. As fatigue varies from person to person, we are interested in knowing what YOU feel. Please answer as many questions as you can.

1. Do you usually feel fatigued?
  a. Yes
  b. No
2. How often have you felt fatigued in the past month? Please tick the answer that best suits you.
  a. Never
  b. Once a month
  c. Once a fortnight
  d. Once a week
  e. 2-5 times a week
  f. Almost every day for a few hours
  g. Every day, all the time
3. Can you predict when you are likely to feel fatigued?
  a. Never, my fatigue just comes on suddenly
  b. Rarely
  c. Sometimes
  d. Often (most times)
  e. All the time
  f. Other, please specify ____________________
4. Do your fatigue levels fluctuate during the day?
  a. Yes
  b. No
5. If yes, when do you usually feel most fatigued? Please tick all that apply to you.
  a. Early morning
  b. Morning
  c. Mid-day
  d. Early afternoon
  e. Afternoon
  f. Evening
  g. Night
6. What words would you use to describe your fatigue? Tick all suitable answers please:
  a. Tiredness
  b. Exhaustion
  c. Malaise
  d. Lethargy
  e. Heaviness
  f. Difficulty concentrating on what I have to do
  g. Low energy
  h. Feeling drained
  i. Other (please specify)______________________________________
7. Have your levels of fatigue increased since your diagnosis of your [Condition]?
  a. I don’t feel fatigued at all
  b. No, my fatigue levels have not changed
  c. Yes, I feel more fatigued since my diagnosis
  d. Fatigue started before I was diagnosed
  e. Not sure
  f. Other _____________________________

#### SECTION 3: Fatigue and how it affects you

The next few questions looks at what sort of activities make you tired.

7. Do activities in your daily routine such as washing, dressing, cleaning, cooking, etc make you fatigued?
  a. Yes
  b. No
8. There may be some routine activities in your daily routine that you have restricted or abandoned, or are planning to restrict <u>because of fatigue</u>.
  a. Which routine activities have you <u>restricted due to fatigue</u>? _______________________________________________________________________________ _______________________________________________________________________________
  b. Which routine activities have you given up <u>due to fatigue</u>? _______________________________________________________________________________ _______________________________________________________________________________
  c. Which activities are you planning to restrict due to fatigue? _______________________________________________________________________________ _______________________________________________________________________________
9. Please tell us about your hobbies or leisure activities _______________________________________________________________________________
10. Have you had to restrict or abandon, or plan to restrict any hobbies or leisure activities due to fatigue?
  a. Yes
  b. No A) Which hobbies or leisure activities have you had to restrict due to fatigue? ___________________________________________________________________________ ___________________________________________________________________________ B) Which hobbies or leisure activities have you had to stop or abandon due to fatigue? ___________________________________________________________________________ ___________________________________________________________________________ C) Which hobbies or leisure activities do you plan to restrict due to fatigue? __________________________________________________________________________ __________________________________________________________________________
11. Is there <u>anything else</u> that triggers your fatigue or makes you feel excessively tired? ___________________________________________________________________________ __________________________________________________________________________ ___________________________________________________________________________ ___________________________________________________________________________ ___________________________________________________________________________
12. If you are in employment (part-time or full time), how <u>does fatigue affect your work</u>? ___________________________________________________________________________ __________________________________________________________________________ __________________________________________________________________________ __________________________________________________________________________ __________________________________________________________________________
13. Does stress affects your levels of fatigue?
  a. All the time
  b. Most of the time
  c. Often
  d. Sometimes
  e. Rarely
  f. Never
14. Does <u>fatigue</u> affect your social life?
  a. All the time
  b. Most of the time
  c. Often
  d. Sometimes
  e. Rarely
  f. Never
15. Does <u>fatigue</u> affect your concentration (e.g. during activities such as driving, reading, etc)
  a. All the time
  b. Most of the time
  c. Often
  d. Sometimes
  e. Rarely
  f. Never
16. Does <u>fatigue</u> affect your **balance**?
  a. All the time
  b. Most of the time
  c. Often
  d. Sometimes
  e. Rarely
  f. Never
17. Does <u>fatigue</u>makes your movements worse?
  a. All the time
  b. Most of the time
  c. Often
  d. Sometimes
  e. Rarely
  f. Never

Thank you for this information. The next section looks at how you cope with fatigue.

**********************************************************************************

#### SECTION 4: Managing Fatigue

18. Have you sought any advice from professionals on how to <u>manage your fatigue</u>?
  a. Yes
  b. No (Please go to question 20)
19. If yes, did you get any advice?
  a. Yes
  b. No A) If yes, **who** gave you advice? ____________________________________ B) **What** advice did you get? ___________________________________________________________ C) **When** did you seek advice? ____________________________________ D) **Did it help**? ______________________
20. **What do you do to help with your tiredness**? Please also include any lifestyle changes, hobbies or leisure activities, or devices that help, if applicable. _______________________________________________________________________________ _______________________________________________________________________________ _______________________________________________________________________________
21. Have you **tried anything to cope with fatigue and <u>founditnottohelp</u>**? _______________________________________________________________________________ _______________________________________________________________________________ _______________________________________________________________________________
22. Do you need to rest during the day to help with your tiredness?
  a. Always
  b. Most of the time
  c. Often
  d. Sometimes
  e. Rarely
  f. Never
23. If yes, for how long do you usually rest? _______________________________________________________________________________
24. Do you still feel tired after taking rest?
  a. Always
  b. Most of the time
  c. Often
  d. Sometimes
  e. Rarely
  f. Never
25. Does exercise help to manage your fatigue levels?
  a. All the time
  b. Most of the time
  c. Often
  d. Sometimes
  e. Rarely
  f. Never
  g. I don’t know
26. On a scale of 1-10, how would you rate your average levels of tiredness in the past month? Please put a cross on the scale below. 1----------------------------------------------------------------------------10 Not tired at all Worst level of exhaustion
27. Please describe anything else that you would like to add about your fatigue not covered in the questions above (what causes it, how it affects you, how you cope, etc). _______________________________________________________________________________ _______________________________________________________________________________

